# Harnessing Mondo and the ILAE Classification for Curation and Analysis of 71,942 epilepsy patient variants

**DOI:** 10.1101/2025.01.07.25320161

**Authors:** Alina Ivaniuk, Tobias Bruenger, Eduardo Perez-Palma, Junhui Xiao, Mark St John, Fábio A. Nascimento, Guo-Qiang Zhang, Samden Lhatoo, Dennis Lal

## Abstract

Mapping epilepsy variants in ClinVar to standard disease classifications remains challenging due to incomplete phenotype information. We integrated the Mondo Disease Ontology (Mondo) with the International League Against Epilepsy (ILAE) Classification to systematically identify, organize, and analyze age-dependent epilepsy-associated variants from the world’s largest public variant repository. Starting from over three million ClinVar entries, we filtered monogenic disorders tagged with 421 Mondo terms representing neonatal/infantile, childhood, or variable-onset epilepsy syndromes. Gene-disease validity was further refined using multiple expert-curated lists, resulting in 71,942 unique variant submissions in 171 genes. Approximately 70% of pathogenic and uncertain variants clustered in 50 genes, highlighting the concentration of clinically relevant findings in a relatively small subset of known epilepsy genes. *SCN1A*, *GRIN2A*, and *DEPDC5* stood out as top contributors to neonatal/infantile, childhood, and variable-onset syndromes, respectively. Notably, two-thirds of genes were specific to a single age group, but the remaining one-third spanned multiple syndromes, reflecting both phenotypic and genetic heterogeneity. We also observed substantial overlap with neurodevelopmental and autism-related genes, especially among early-onset epilepsies, emphasizing shared molecular pathways. Our Mondo-driven approach demonstrates the feasibility of large-scale, ontology-based curation of ClinVar for domain-specific analyses, offering greater clinical precision than traditional text-based queries. This resource can guide epilepsy-focused gene panel design, inform new classification updates, and pave the way for age-targeted therapeutics. By bridging the gap between extensive genomic repositories and detailed clinical ontologies, we provide a robust, standardized framework to accelerate progress in precision epilepsy genetics.

## Introduction

Epilepsy is one of the most common neurological disorders that affects over 45 million people worldwide and displays a striking heterogeneity in etiologies and clinical presentations.^1^ Genetic etiologies of epilepsy are becoming increasingly recognized, and the number of genes associated with monogenic epilepsies continues to grow. A recent curation effort study identified over 700 epilepsy-related genes through automated literature mining and manual expert curation, while another integrated genetic panel data from large laboratories with automated searches and manual review to expand this list to over 900 genes.^2,3^ Despite the extensive list, studies based on real-world cohorts show a disproportional representation of a limited set of genes associated with early-onset epilepsies, such as *SCN1A*, *KCNQ2*, and *CDKL5*.^4–7^ These early-onset epilepsy syndromes are characterized by distinct, oftentimes severe, phenotypes with comorbid neurodevelopmental delay (NDD) and behavioral disorders.^8–10^ Moreover, increasing evidence suggests an overlap between genes associated with epilepsies, NDD, and behavioral disorders such as autism.^10–12^ However, while early seizure onset is strongly associated with genetic etiology^13,14^, emerging qualitative evidence suggests a distinct genetic landscape in later-onset syndromes, including childhood and adult-onset epilepsies, with a broader phenotype spectrum including focal lesional and non-lesional epilepsies. ^6,12,15–17^ Yet, the bias in referrals for genetic testing in early age^6^ hinders a holistic appraisal of later-onset epilepsy syndromes in real-world cohort studies.

As rare disease genomics, phenotypic spectra, and populations of interest in epilepsy expand, the ability to integrate and interpret rapidly growing genomic datasets in a clinically meaningful manner is becoming increasingly critical. Standardized, ontology-driven frameworks offer solutions for organizing and analyzing complex genotype-phenotype relationships, thereby streamlining variant classification, promoting data consistency, and accelerating the translation of genomic insights into clinical care.^18,19^ While ontologies such as Mondo Disease Ontology (Mondo) and the Human Phenotype Ontology (HPO) have advanced our capacity to harmonize terminology across studies ^18,20,21^, effectively leveraging them within large-scale genetic variant repositories remains challenging. Databases like ClinVar, which aggregate millions of user-submitted variant interpretations^22^, are the largest resource of patient variants – including epilepsies. However, this genetic data-rich resource lacks robust phenotype-to-genotype mapping that aligns with epilepsy classification systems that would make classifications more accessible for clinicians. Using the current ClinVar front-end interface, a user may search for epilepsy-associated variants by directly supplying the term “epilepsy,” yet it yields unstructured results with no opportunities for filtering out noise. Many epilepsy syndromes don’t contain the term ‘epilepsy’ in their name (e.g., Dravet Syndrome, West Syndrome, Developmental Epileptic Encephalopathy, etc.) and are not identified, aggregated, and displayed when searching for ‘epilepsy.’ Several web platforms improving the ClinVar user experience have been developed by the community^23,24^, but the issue of phenotype-driven ClinVar data filtering is yet to be addressed.

Addressing the limitations of ClinVar for phenotype-specific analyses requires a standardized framework that not only harnesses disease ontologies but also integrates clinically meaningful classifications, ultimately allowing for systematic filtering. In the epilepsy domain, the Classification and Definition of Epilepsy Syndromes of the International League Against Epilepsy (further referred to as ILAE Classification) represents the reference epilepsy classification developed by the International League Against Epilepsy (ILAE) Nosology and Definitions Task Force for use by healthcare professionals, researchers, policymakers, and other stakeholders across the globe.^25^ The ILAE Classification is particularly focused on age-dependent epilepsy syndromes (ILAE-ADES), reflecting accumulated research and empiric evidence about the age of onset of particular epilepsy syndromes.^26–28^ It distinguished syndromes with Onset at Neonatal/Infantile, Childhood, and Variable Age—each with distinct clinical features, etiologies, including genetic factors, and prognoses.^26–28^ On the other hand, ClinVar variant submissions are supplied with Mondo^21,29^, a hierarchical disease-specific ontology that can be leveraged for structuring the phenotypic data embedded in the database. Integrating Mondo-based annotations with ILAE Classification has the potential to filter ClinVar more precisely, enabling the construction of large, clinically relevant, and understandable to clinicians’ gene and variant sets.

Here, we describe a scalable, ontology-based filtering framework that bridges ClinVar’s genetic data with the ILAE-defined classification of epilepsy syndromes. By mapping over 600 Mondo terms to ILAE-ADES, we refined more than three million ClinVar variant submissions to identify a focused dataset of 171 genes and 71,942 variants—constituting the largest, most clinically aligned epilepsy-specific genetic resource assembled to date. To ensure data reliability and mitigate biases, we incorporated a multi-tiered gene-disease confidence framework informed by multiple expert-curated resources. The subsequent analyses highlight genes associated with ILAE-ADES, examine overlaps in gene sets between ILAE-ADES, neurodevelopmental disorders (NDDs), and autism spectrum disorder (ASD), and provide proportions of genes associated with lesional epilepsies. Our study offers insights for refining diagnostic gene panels, provides data for guiding clinical resource allocation, education in clinical genetics of epilepsy, and guidelines and policy making, and demonstrates the application of systematic ontology-driven methods for disorder-specific genomic research.

## Methods

### Overview of Approach to Dataset Acquisition

We developed a framework to extract epilepsy-specific variants from ClinVar by leveraging embedded variant phenotype annotations linked to Mondo Disease Ontology terms. These terms were curated and mapped to the terms of the 2022 International League Against Epilepsy (ILAE) Classification of Epilepsy (further referred to as ILAE Classification).^26–28^ Using this framework, we identified a subset of genes and variants associated with ILAE-defined age-dependent epilepsy syndromes (ILAE-ADES). A detailed workflow summarizing the process is shown in Figure 1A, with additional details on each individual step provided in the Supplemental Methods.

**Figure 1.**
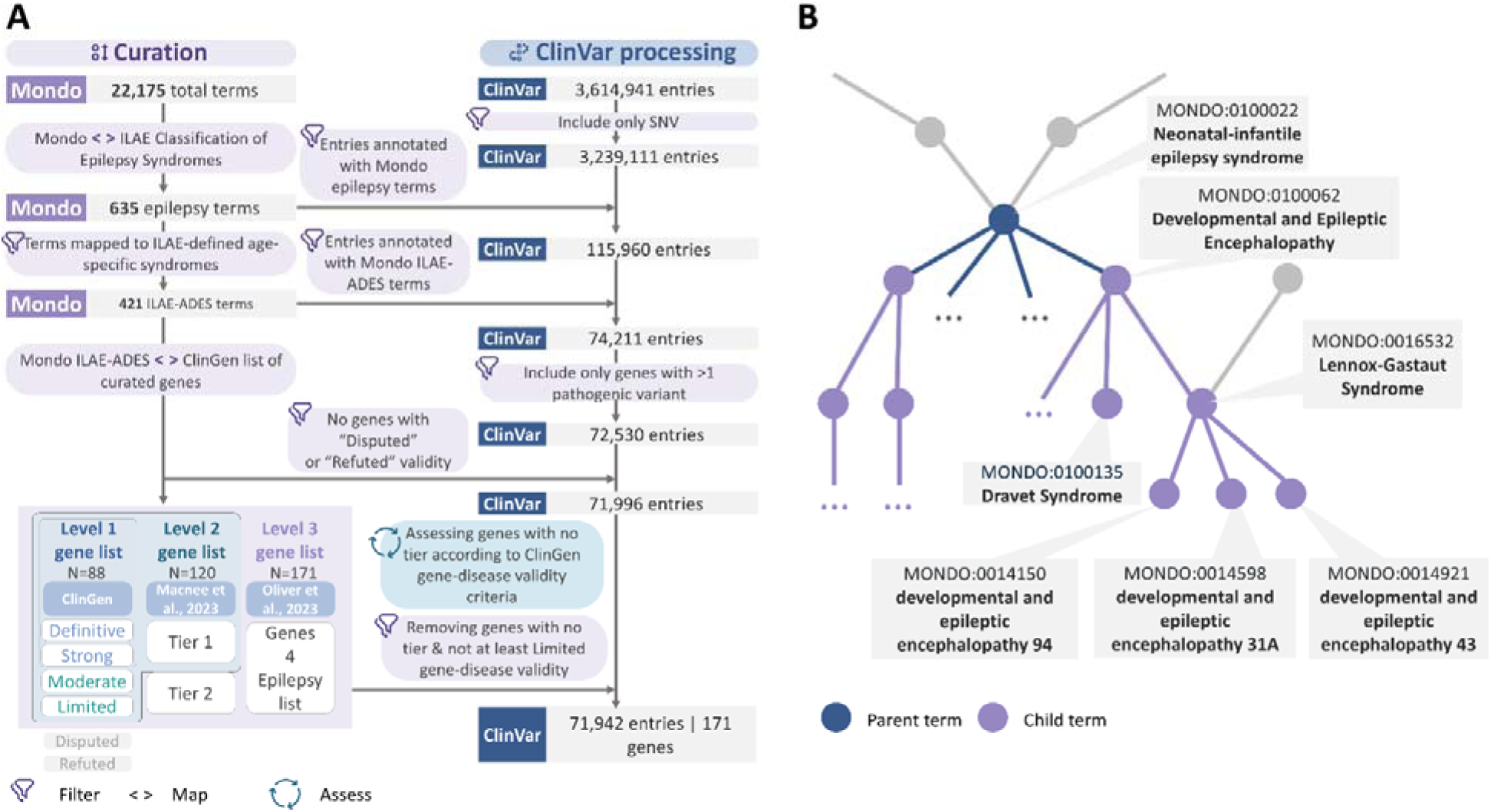
A. Mondo terms mapping and ClinVar dataset processing. We cross-mapped the Mondo Disease Ontology terms with ILAE Classification of Epilepsy Syndromes to derive 635 terms associated with broad epilepsy classification concepts and 421 terms associated with the ILAE-ADES. We used this dictionary to filter over 3 million ClinVar variant submissions to identify 71,942 variant entries linked to 171 genes associated with ILAE-ADES. The genes and variants were further validated through stringent criteria, including gene-disease association evidence and tiered stratification based on clinical relevance. (ClinGen curation lists, epilepsy gene tiers from Macnee et al. 2023, and gene list from Oliver et al., 2023). B. Illustration of hierarchical Mondo structure. We identified most non-specific terms mapped to the ILAE Classification (“parents”) and included them, as well as all terms downstream of them (“children”), to identify a set of terms associated with ILAE-ADES. Abbreviations: ILAE-ADES, ILAE-defined age-dependent epilepsy syndromes

To address the limitations of phenotype-guided filtering in ClinVar, we utilized Mondo’s hierarchical disease ontology, which integrates terms from 18 biomedical ontologies to enable cross-referencing and quantitative analysis of healthcare data.^21^ Of the 3,614,941 variant submissions and 3,173,133 unique variants available in ClinVar, 1,690,706 (46.8%) variant submissions and 1,455,371 (45.9%) unique variants were annotated with Mondo terms, enabling a pilot of scalable and reproducible filtering strategy.

### Mapping Mondo Terms to the ILAE Classification of Epilepsy

We mapped Mondo terms to the ILAE Classification to enable ontology-based filtering specific to epilepsy phenotypes. The ILAE Classification defines epilepsy syndromes by age of onset, clinical features, etiologies, and prognoses. The complete Mondo ontology (v2022-03-01) was downloaded from the Monarch Initiative GitHub repository, and ILAE-defined epilepsy concepts were accessed from EpilepsyDiagnosis.org and related ILAE publications.^26–28^

Each ILAE epilepsy concept was individually assessed and manually mapped to corresponding Mondo terms, including parent and child terms (see Supplemental Table 1 for parent terms). We refined these mappings to focus on ILAE-ADES, categorizing Mondo terms into three onset categories according to the ILAE Classification: Neonatal/Infantile, Childhood, and Variable Age. Clinical descriptions in the ILAE Classification were used to annotate lesional epilepsy syndromes and corresponding genes. Syndromes resulting in non-specific anatomical changes (e.g., atrophy) were considered non-lesional. Details on mapping criteria and derived terms are provided in Supplemental Table 2.

Deriving Genes and Variants Associated with ILAE-Defined Mondo Terms in ClinVar The complete ClinVar public dataset was accessed and downloaded from the FTP site (https://ftp.ncbi.nlm.nih.gov/pub/clinvar/xml/) in an XML format on December 7, 2023, and converted to a tabular format. We filtered the ClinVar dataset to include entries annotated with the identified epilepsy-specific Mondo terms and further narrowed it to ILAE-ADES terms.

Variants were restricted to single-nucleotide variants (SNVs) associated with monogenic epilepsy disorders. We excluded genes without known pathogenic or likely pathogenic (P/LP) variants to ensure reliability and reduce noise.

To facilitate the quality control of the derived dataset, ensure the reliability of our findings, and set our list in the context of existing expert-driven curation efforts, we assigned three levels of gene-disease validity confidence to the resulting set of genes. The levels were based on the three most comprehensive curation projects: ClinGen, Macnee et al., and Oliver et al. ^2,3,30^ The levels were arbitrarily derived based on the methodology employed by curators and the broadness of the resulting gene lists. As the strongest evidence of gene-disease association, we used curations by ClinGen, a collaborative expert-driven program supported by the US National Institute of Health that synthesizes information to aid the clinical use of genetic information and develops tools to facilitate the widespread use of this information.^31^

- Level 1: Definitive and Strong gene-disease associations per ClinGen.
- Level 2: Moderate or Limited validity per ClinGen and Tier 1 genes from Macnee et al.
- Level 3: Tier 2 genes from Macnee et al. and genes from Oliver et al. not included in Levels 1 or 2.

Genes classified as “Disputed” or “Refuted” by ClinGen were excluded. Two genes (*ARSD* and *SLC28A2*) were excluded after manual review due to insufficient evidence of gene-disease validity (see Supplemental Results).

### Additional Gene Lists for Comparison

There is increasing evidence of monogenic etiologies underlying neurodevelopmental disorders (NDDs) and autism spectrum disorder (ASD).^32,33^ NDD, ASD, and epilepsy are often combined within a phenotypic spectrum of the same disorder, and new data on shared genetic landscapes of these disorders is emerging.^10–12^ We evaluated the overlap between ILAE-ADES-associated genes and those implicated in NDD and ASD. NDD genes were sourced from the SysNDD database^34^, excluding entries marked as “Refuted” or “Not applicable.” ASD-associated genes were retrieved from the SFARI Gene database^35^ and filtered to include high-confidence (score 1) and strong candidate (score 2) genes.

### Statistical Analysis

We used descriptive statistics to summarize categorical data. Fisher’s exact test and Wilcoxon rank-sum test were employed for comparisons of categorical and continuous variables, respectively, with false discovery rate (FDR) correction for multiple testing. A significance level of p=0.05 was applied. Statistical analysis and visualization were performed using R (v4.4.0) in RStudio (v2022.12.0).

## Results

### Integration Of Large-Scale Genomic Data from ClinVar With Clinical Grade Epilepsy Syndromes

Epilepsy’s clinical and genetic heterogeneity complicates identifying genetically and clinically homogeneous cohorts for large-scale research. A significant challenge is the lack of standardized clinical coding aligned with international frameworks, limiting precision in biobank-scale studies. While ClinVar, the largest repository of epilepsy-associated genetic variants, contains extensive data, it has yet to be fully utilized for syndrome-specific genetic landscape studies. This limitation stems from the absence of ILAE-grade classification systems and ontology-based filtering tools.

To address these gaps, we leveraged ClinVar’s use of the Mondo Disease Ontology for disease annotation. By mapping Mondo terms to ILAE-defined epilepsy syndromes (as described in the Methods section on “Mapping Mondo terms to the ILAE Classification of Epilepsy”), we extracted epilepsy-relevant variants and refined these into subsets specific to age-dependent epilepsy syndromes (ILAE-ADES). This process enabled a more focused analysis of epilepsy-associated genetic variants in ClinVar, facilitating detailed phenotype-genotype studies. As an initial step, we performed a mapping of Mondo ontology terms to ILAE-defined epilepsy classifications and the ILAE-defined age-dependent epilepsy syndromes (ILAE-ADES, see Methods). This mapping process identified 635 Mondo terms linked to ILAE epilepsy concepts, including 421 terms associated explicitly with ILAE-ADES categories (Figure 1), thereby facilitating the derivation of more granular and clinically relevant variant datasets. Of 3,173,133 ClinVar entries, we derived a subset of 115,960 monogenic variant submissions associated with at least one of 635 Mondo terms for ILAE-defined epilepsy concepts and further filtered it to 74,211 submissions associated with 421 Mondo terms associated with ILAE-ADES (see Methods, Figure 1).

### Large-scale curation of variant submissions and gene-disease validity

Before downstream analyses of the clinical genetic spectrum of epilepsies according to ILAE classification, we developed a conservative approach to remove genes with limited gene-disease validity. First, we only included genes with at least one pathogenic variant to remove genes with no or limited association with epilepsy, resulting in 72,530 entries in a total of 175 genes. To further increase the validity of the epilepsy-ILAE gene set, we excluded gene-phenotype pairs with ‘Disputed’ or ‘Refuted’ gene-disease validity according to the ClinGen consensus gene-disease validity expert group, yielding 71,996 entries variant submissions for 173 genes. Since our approach is the largest genotype-phenotype in epilepsy thus far, we performed further gene-disease validity assessments to focus only on well-established epilepsy-associated genes utilizing additional manually curated resources from domain experts (see Methods for details). Two genes, *ARSD* and *SLC28A2*, were removed (see Supplemental Results). The final data set of genes associated with ILAE-ADES consisted of 171 genes encompassing 71,942 variant submissions and 61,548 unique variants.

### Comprehensive Analysis of Pathogenic and VUS Variants in ILAE-Defined Epilepsy Genes: Insights into Gene-Disease Associations and Submission Trends

Next, we explored the genetic spectrum across ILAE-defined epilepsy syndromes. We conducted further analysis accounting for P/LP variants, which imply genetic diagnosis, and VUS, which can potentially gain clinical relevance as reclassification opportunities arise. Of the 40,324 variant submissions associated with ILAE-ADES, 34,300 were unique variants, indicating that approximately 15% of submissions were for likely recurrent variants classified as P/LP or VUS (Table 1). Notably, over 70% of variant submissions (N=28,687) and unique variants (N=24,103) were linked to just 50 genes (Figure 2A). Among these, *TSC2* had the highest number of P/LP and VUS submissions (N=4,268, 10.5% of P/LP and VUS submissions), while *SCN1A* had the most P/LP variant submissions (N=1,350, 17% of all pathogenic variant submissions). Additionally, 111 genes (65%) associated with ILAE-ADES overlapped with genes implicated in NDD, and 49 genes (28.6%) were also linked to both autism and NDD (Figure 2B).

**Figure 2.**
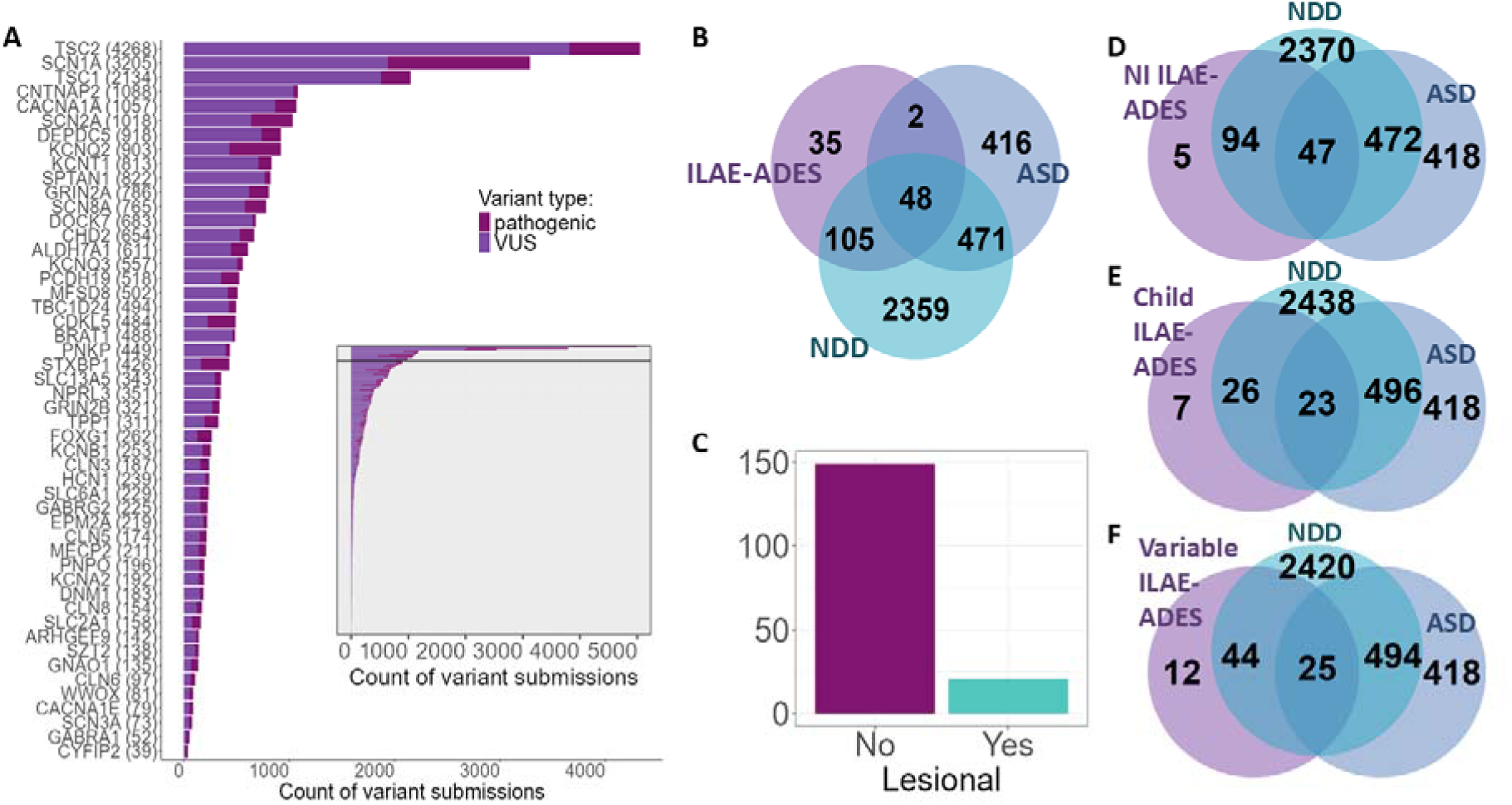
A. Fifty Mondo-defined epilepsy genes with the highest number of patient submissions for pathogenic variants and VUS. The numbers in brackets next to the gene names indicate the total number of submissions, and the color designates the clinical significance of the variants. The pane on the right side illustrates the variant count per gene in all 171 genes. Forty-three genes without an asterisk belong to Level 1, and five genes with a single asterisk are genes from Level 2. A pane in the right part of the plot shows variant count in all 171 included ILAE-defined age-dependent epilepsy genes. The horizontal black line indicates the 50 genes visualized in pane A, which host over 80% of all variants and over 70% of patient submissions associated with age-dependent epilepsy syndromes in this study. B. A Venn diagram of the overlap of genes associated with ILAE-defined age-dependent epilepsy syndromes and genes associated with neurodevelopmental disorders (NDD) and autism. Sixty-four percent of epilepsy genes in the study overlapped with NDD genes, and 27% - with genes causing autism and NDD. C. The counts of age-dependent epilepsy genes associated with lesional and non-lesional epilepsy. Most of the genes (87%) were associated with non-lesional epilepsy. D-F. Venn diagrams of gene sets overlap in three ILAE-ADES categories (neonatal/infantile, childhood, and variable onset in frames D, E, and F, respectively) with gene sets for NDD and ASD. We observed a decrease in the proportion of the overlap from the early-onset neonatal/infantile category towards the variable onset category, in which epilepsy can manifest in late teenage to adulthood (p=0.003). Abbreviations: ASD, autism spectrum disorder; Child, childhood-onset; ILAE-ADES, ILAE-defined age-dependent epilepsy syndromes; NDD, neurodevelopmental disorders; NI, neonatal/infantile-onset; Variable, variable-onset.

**Table 1.**
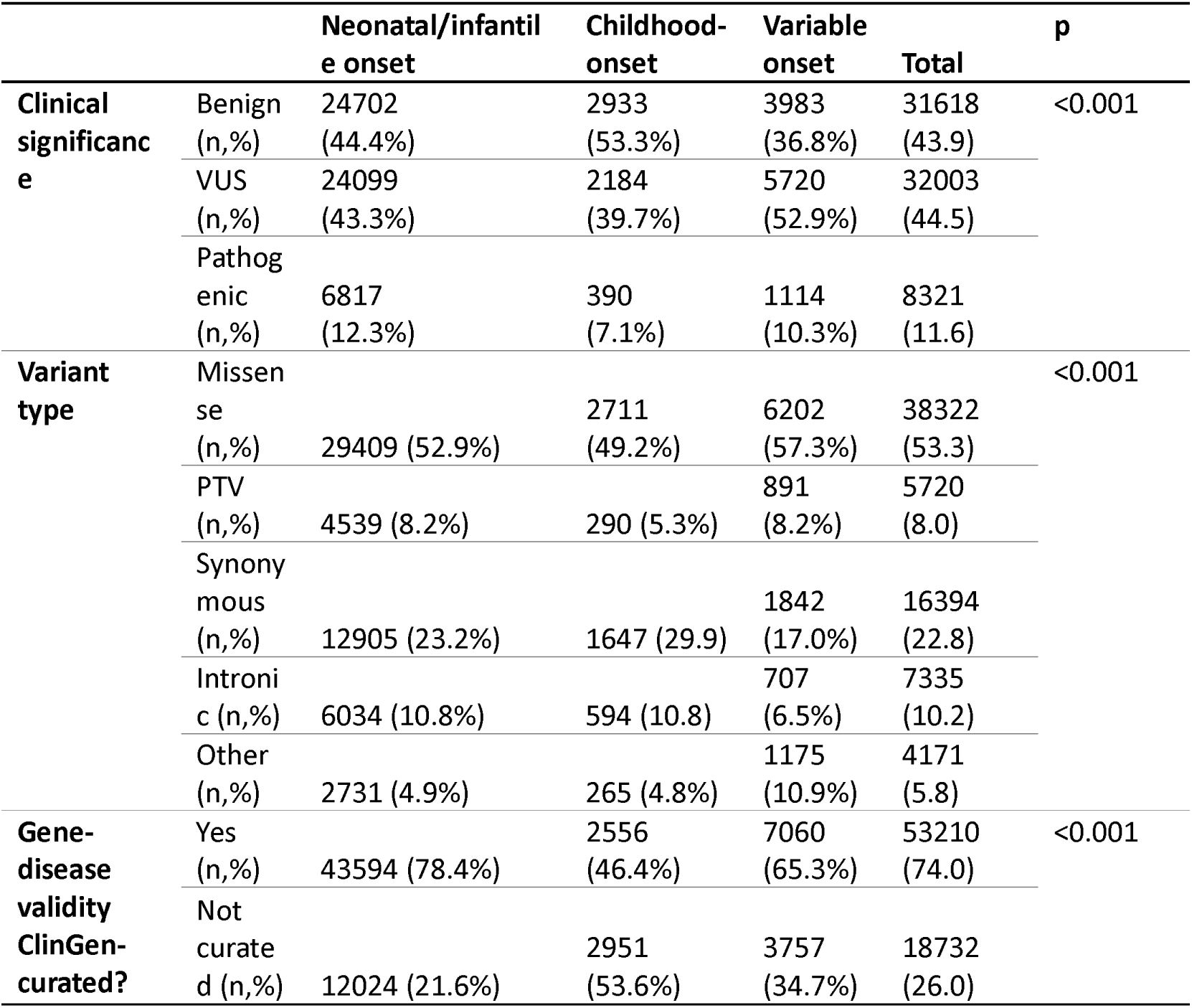
Summary of clinical significance, variant type, and gene-disease validity curation status of 71,942 variant submissions across all ILAE-defined age-dependent epilepsy syndromes groups. The last row shows the number of submissions for which the associated gene-phenotype pair was curated by ClinGen. Abbreviations: PTV, protein-truncating variant.

Notably, of 146 genes associated with neonatal/infantile ILAE-ADES, 141 (96.6%) overlapped with the genes causing NDD, ASD, or both, compared to 49 of 56 (87.5%) in the childhood ILAE-ADES and 69 of 81 (85.2%) in the variable ILAE-ADES (p=0.003; Figure 2 D-F). Most genes (N=164, 87%) were related to non-lesional epilepsy (Figure 2C) compared to 13% of genes associated with epilepsy syndromes associated with epileptic lesions.

ClinVar serves as a historical resource, making cross-sectional analyses of its data prone to historical diagnostic biases. To evaluate the extent of bias and examine temporal trends, we compared variants submitted to ClinVar over the last two years with those submitted in all previous years. Over this recent two-year period, pathogenic variants were reported for 158 ILAE-ADES genes. Notably, the 50 genes with the highest number of epilepsy-related submissions remained consistent between the recent and historical datasets, suggesting stability in gene prioritization over time. Additionally, we observed a strong correlation between the total number of submissions per gene and age category in the two-year dataset and the cumulative dataset (r = 0.77; 95% CI: 0.7–0.82, p < 2.2 × 10⁻¹). These findings indicate that while temporal trends influence data collection, the core set of genes associated with epilepsy syndromes has remained largely unchanged (Figures S3 and S4).

### A massive scale investigation of the genetic landscape of ILAE-defined age-dependent epilepsy syndromes

The International League Against Epilepsy (ILAE) classifies age-dependent epilepsy syndromes into three distinct categories: Onset in Neonates and Infants, Onset in Childhood, Onset at Variable Age (designated Neonatal/infantile, Childhood, and Variable onset in our analysis).

Missense variants represented the majority across all variants associated with ILAE-ADES (52.9%-57.3%), with protein-truncating variants and synonymous variants contributing less frequently, highlighting consistent patterns of variant types in genetic epilepsy across age categories (Table 1). The majority (64.91%; 111/171) of genes were associated with P/LP variants in a single age category, while 60 genes were associated with multiple age-related categories (Figure 3A). Notably, no gene was associated with all three age-related categories when focusing on genes where pathogenic variant submissions accounted for more than 20% of the total submissions (Figure 3B). Most of the genes were linked to non-lesional epilepsy, and the variable onset category had the highest proportion (16.07%; 13/81) of genes associated with lesional epilepsy (Figure 3C).

**Figure 3.**
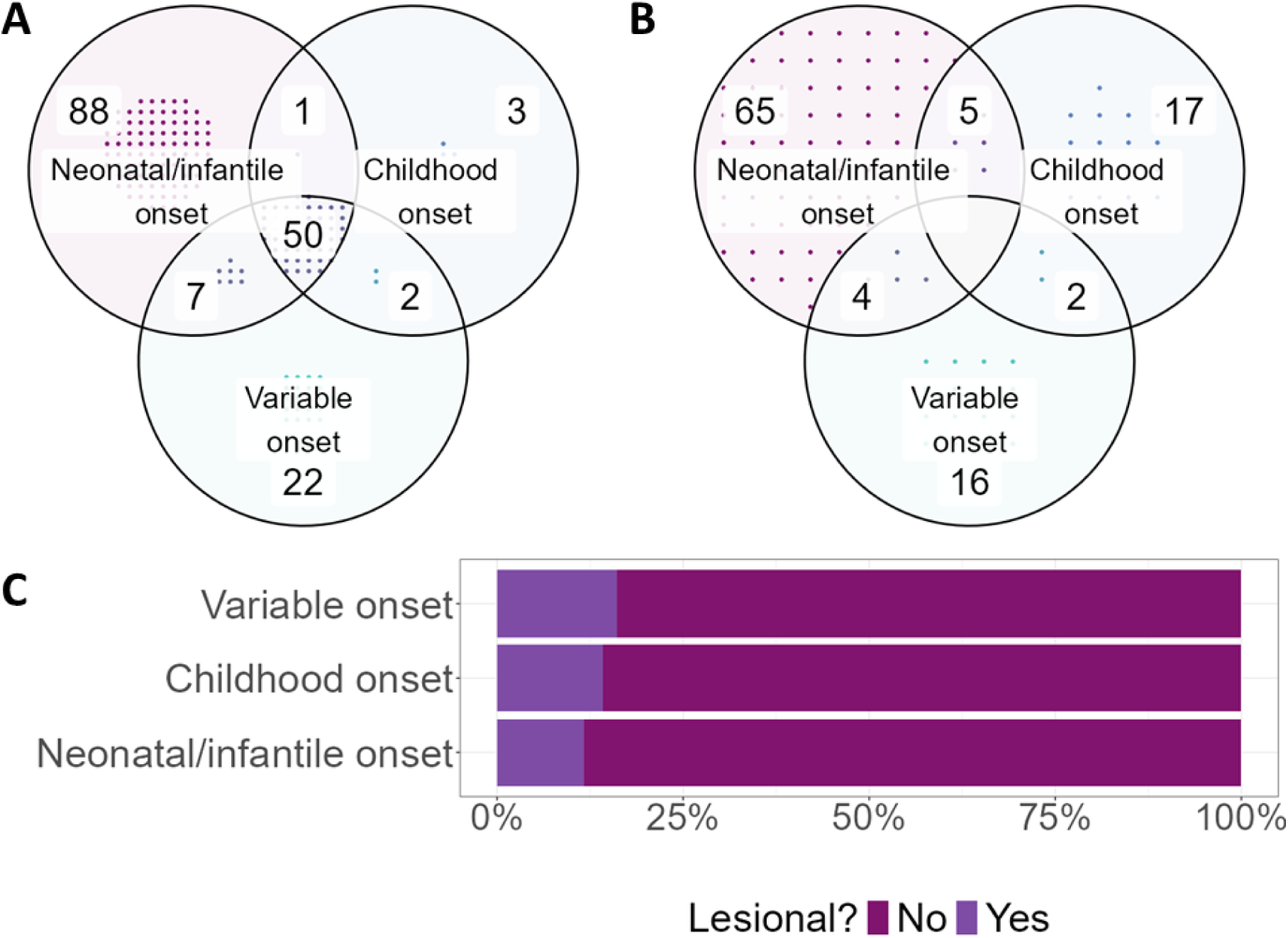
A-B. Density Venn diagrams showing an overlap between genes in different age categories. Frame A shows overlap between age categories in all included genes. Frame B shows overlap of age categories with genes for which pathogenic variants constitute at least 20% of all variant entries. Higher ratio of pathogenic variants results in a higher age-dependent specificity of a gene. C. A barplot showing the proportions of genes causing lesional epilepsy per age category. Lesional epilepsy syndromes constituted the minority of all genes in all categories, with an increasing ratio of lesional epilepsies in age categories with later onset.

The neonatal/infantile epilepsy syndromes category was the most represented among age-dependent syndrome groups, with 146 genes linked to 55,618 variant submissions, including 6,817 pathogenic variants (12.25%) (Figure 4A). Fifteen genes had over 80 pathogenic variants submissions, including *SCN1A* (N=1350), *TSC2* (N=670) *KCNQ2* (N=486), *SCN2A* (N=390), *TSC1* (N=283), *STXBP1* (N=275), *CDKL5* (N=268), *SCN8A* (N=200), *CACNA1A* (N=199), *PCDH19* (N=170), *ALDH7A1* (N=163), *FOXG1* (N=138), *CHD2* (N=131), *SLC2A1* (N=86), and *KCNB1* (N=81). Developmental and epileptic encephalopathies were the predominant phenotype group (62.89% of pathogenic submissions) in this age category (Figure 4B). Childhood-onset epilepsy syndromes were associated with a total of 56 genes with 5,507 patient submissions (390 (7.1 %) pathogenic). Three genes accounted for over 80% of all pathogenic variant submissions in this category (Figure 4C): *GRIN2A* (N=187), *SLC6A1* (N=76), and *KCNT1* (N=71), reported to be associated with Landau-Kleffner syndrome, myoclonic-astatic epilepsy, and autosomal dominant nocturnal frontal lobe epilepsy, respectively. Epilepsy syndromes with variable onset were associated with 81 genes and 10,817 submitted variants (1114 (10.3%) pathogenic). Six genes had over 50 pathogenic variant submissions (Figure 4E): *DEPDC5* (N=185) and *NPRL3* (N=51) associated with familial focal epilepsy with variable foci, and *TPP1* (N=133), *MFSD8* (N=92), *CLN3* (N=87), and *CLN5* (N=66) associated with progressive myoclonic epilepsies.

**Figure 4.**
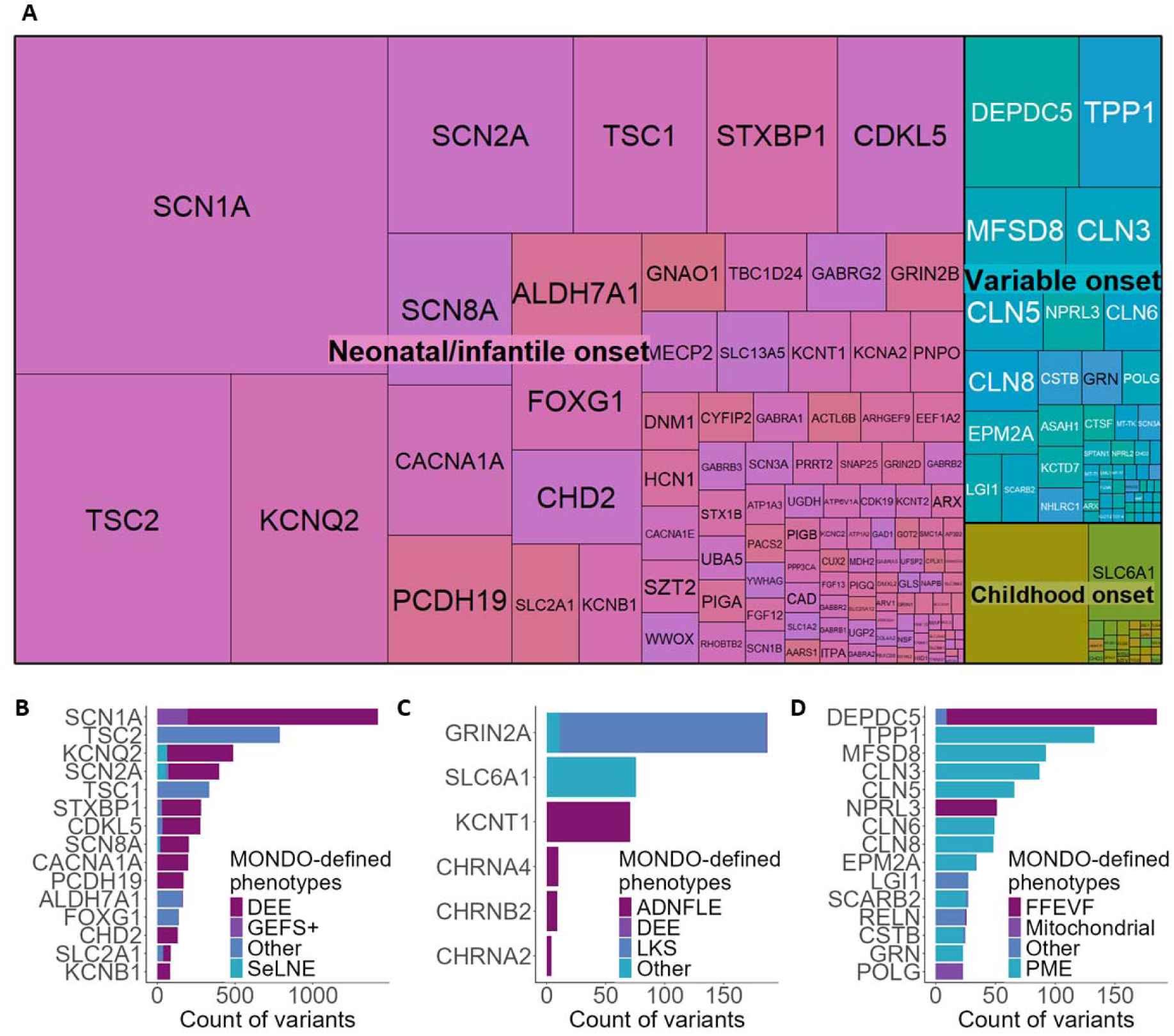
Overview of genes associated with Mondo-defined age-dependent epilepsy syndromes. Genes with at least five submissions of pathogenic variants are displayed in all the frames. A. A treeplot showing the gene-wise proportion of pathogenic variant submissions across age-dependent categories. B-D. Bar plots showing pathogenic variant submission counts for genes with the highest number of pathogenic variant submissions associated with Mondo-defined epilepsy syndromes with neonatal/infantile (B), childhood (C), and variable (D) onset. The color codes for the Mondo phenotypes. Number- or gene-denoted phenotypes were aggregated into higher (more unspecific) Mondo hierarchy (for example, specific Mondo terms like developmental and epileptic encephalopathy, 42 (MONDO:0014917) and CDKL5 disorder (MONDO:0100039) were aggregated into developmental and epileptic encephalopathy (MONDO:0100062)). Abbreviations: ADNFLE, Autosomal dominant nocturnal frontal lobe epilepsy; DEE, Developmental and epileptic encephalopathy; FFEVF, Familial focal epilepsy with variable foci; GEFS+, Generalized epilepsy with febrile seizures plus; LKS, Landau-Kleiffner syndrome; PME, Progressive myoclonic epilepsy; SeLNE, Self-limited neonatal (infantile) epilepsy.

## Discussion

We present the largest investigation of the genetic landscape of epilepsy to date. Our analysis underscores both the complexity and promise of large-scale, ontology-based interrogation of genetic data in epilepsy. By coupling ClinVar variant submissions with an ontology-driven approach founded on ILAE-defined syndromes, we were able to systematically parse a massive genetic repository and characterize syndrome-specific gene and variant architectures. Notably, our classification pipeline resolved over 71,000 variant submissions into three ILAE age-dependent epilepsy categories, capturing both canonical genes (e.g., *SCN1A*, *KCNQ2*, *STXBP1*) and emerging contributors, while clarifying overlaps with neurodevelopmental and autism-associated genes. The high rate of shared genes among neonatal/infantile epilepsies and NDD/ASD highlights the deep interconnectivity between epilepsy subtypes and broader neurodevelopmental phenotypes.^10–12^ In parallel, the enrichment of gene-disease associations in established syndromes such as familial focal epilepsy or progressive myoclonic epilepsies demonstrates how systematic ontology mapping can unmask population-scale trends. Collectively, these observations provide robust evidence that leveraging disease ontologies like Mondo, in tandem with expert-based frameworks such as the ILAE Classification, can refine our understanding of genotype-phenotype spectra in epilepsy.

Mondo is a structured ontology for human diseases encompassing over 15,000 terms for rare disorders, making it especially attractive for genetic conditions research.^21^ Mondo is widely used for ontology-based disease modeling and multidomain rare disease data harmonization. For example, the Kidney Precision Medicine Project incorporated Mondo to harmonize disease definitions across various data sources to develop Kidney Tissue Atlas, a resource that integrated clinical data, tissue samples, and advanced molecular profiling to support precision medicine in renal disease.^36^ Similar Mondo applications to medical domain-specific disease models exist in other domains, such as toxicology and environmental medicine.^37,38^ To our knowledge, our study represents the first application of Mondo to epilepsy genetics research overall and to manipulate ClinVar data to derive the relevant subset of data in particular. The National Center for Biotechnology Information has updated its submission processes for ClinVar and the Genetic Testing Registry to accept Mondo IDs in 2020.^29^ Considering the recentness of this integration, other efforts to leverage it are likely yet to emerge. The modernity of Mondo addition also explains that under 50% of all ClinVar submissions were annotated with Mondo terms. Our study proves the feasibility of using Mondo for disease-specific ClinVar data utilization, which would encourage continued integration of Mondo into ClinVar and promote more submissions supplied with Mondo to enhance the consistency and utility of genetic variant data.

We could directly cross-map every concept in the most recent ILAE Classification to Mondo terms, suggesting that epilepsy concepts are well-represented in Mondo. Such comprehensive coverage of epilepsy terms in Mondo, including terms for rare epilepsies, aligns with expectations given the ontology was constructed semi-automatically from 18 external ontologies and dictionaries, including Human Phenotype Ontology (HPO), in which the ILAE Epilepsiome Task Force curated the seizure sub-ontology according to the 2017 ILAE Classification of Seizure Types. ^20,39–41^ We noted that upper-level, less specific (parent) Mondo terms correspond well to the 2022 ILAE Classification of Epilepsy Syndromes. Yet we also observed several obsolete (e.g., MONDO:0100030 “adolescent/adult-onset epilepsy syndrome,” which earlier corresponded to the ILAE Classification’s group of syndromes with Onset in Adolescents/Adults and now are part of syndromes with Onset in Variable Age) and redundant (e.g., MONDO:0010898 “autosomal dominant epilepsy with auditory features” and MONDO:0100031 “adolescent/adult-onset autosomal dominant epilepsy with auditory features”) categories, which did not impede analysis but present an opportunity for ontology improvement. This redundancy is to be expected given the number of resources used for Mondo creation. According to the Mondo creators’ analysis, over 6,000 out of 10,393 terms for rare disorders were present in three or more sources from which the ontology was constructed.^42^ Enhancing harmonization between Mondo and the ILAE Classification through dynamic expert curation driven by the user community could further increase the utility of Mondo as a tool for epilepsy research.^43^

Using Mondo-based ClinVar filtering and ascertaining the dataset according to the recent epilepsy gene curation efforts, we identified 171 genes associated with ILAE-ADES. Notably, nearly three-quarters of all variants were attributed to just 50 genes. This finding aligns with prior studies evaluating the utility of clinical genetic panels in epilepsy, which demonstrate that pathogenic variants predominantly cluster within a subset of genes despite the expansion of diagnostic panels.^6,7,44^ The 171 genes identified in our analysis show strong concordance with findings from previous studies investigating the genetic landscape of epilepsy. For example, in a recent analysis by the Epi25 collaboration, protein-truncating variants in *NEXMIF*, *SCN1A*, *SYNGAP1*, *STX1B*, and *WDR45* reached exome-wide significance for association with epilepsy.^45^ Of these, only *WDR45* is absent from our gene list. This gene is clinically associated with β-propeller protein-associated neurodegeneration (BPAN), a subtype of brain iron accumulation syndromes characterized by a neurodevelopmental disorder with early-life seizures that typically resolve with age.^46^ The *WDR45*-related disorder is missing in 2022 ILAE Classification, which guided our mapping and subsequent ClinVar filtering, and therefore the gene was not included in the list.

Similar considerations explain differences between our identified age of onset-associated genes lists and those reported in the study by McKnight et al., the until now largest investigation clinical genetic testing study across various ages of epilepsy onset.^6^ While their findings for infantile and early childhood groups align closely with our neonatal/infantile ILAE-ADES category, and their adolescence and adult-onset groups overlap with our variable onset gene set. These differences likely arise from different methodological approach. McKnight et al. categorized genes based on the actual age of epilepsy onset in participants, whereas our study strictly adheres to the 2022 ILAE Classification and Definitions of Epilepsy Syndromes. For example, genes like *TSC2*, commonly associated with infantile spasms, are included in our neonatal/infantile category as defined by the ILAE Classification, despite their potential variable onset. Conversely, genes linked to brain malformations, such as *FLNA* and *PAFAH1B1*, are excluded from our gene sets due to their absence in the ILAE-defined syndromic categories.

While recording the actual age of onset in patients may be more effective for identifying gene-level patterns of epilepsy onset, the ILAE syndrome classification accounts for associated comorbidities. Our mapping approach, based on this classification, provides the only feasible method for leveraging ClinVar, the largest genetic data repository, for research purposes.

The substantial overlap observed between ILAE-ADES genes and those implicated in NDD and ASD highlights shared neurodevelopmental pathways influencing both seizure susceptibility and broader cognitive and behavioral phenotypes.^47^ Notably, nearly 90% of neonatal/infantile ILAE-ADES genes were also implicated in NDDs, and over 28% overlapped with both NDD and ASD-associated genes.^47,48^ This convergence underscores the interconnectedness of epileptic syndromes with broader neurodevelopmental conditions, likely driven by genes critical for synaptic function, ion channel regulation, and neurodevelopmental signaling pathways.

Furthermore, genes such as *SCN1A*, *STXBP1*, and *SYNGAP1* exemplify the dual role of genetic variants in driving epileptic phenotypes and contributing to intellectual disability and autism-related traits.^47^ The enriched overlap in early-onset syndromes suggests a heightened vulnerability during critical periods of brain development, reinforcing the need for age-specific therapeutic strategies.^47^ These findings also advocate for the incorporation of broader neurodevelopmental phenotypic data into epilepsy-focused ontological frameworks, enabling more refined genotype-phenotype analyses and facilitating the discovery of targeted interventions.^49^ Future work should explore whether shared genetic etiologies predict treatment response across these overlapping domains, paving the way for integrative precision medicine approaches.^47,48^

The predominance of non-lesional epilepsy genes in our dataset, representing 87% of the identified genes, highlights the limitations of current diagnostic modalities that focus on germline variants detectable through commercial genetic testing platforms. Lesional epilepsies, often linked to malformations of cortical development (MCD) such as focal cortical dysplasia (FCD), are increasingly associated with somatic mutations in critical pathways like mTOR, which remain undetected in standard germline testing approaches.^50^ For example, somatic variants in mTOR pathway genes, such as *DEPDC5*, *TSC1*, and *TSC2*, have been implicated in FCD and hemimegalencephaly, emphasizing their role in structural brain abnormalities and epilepsy pathogenesis.^51,52^ Recent studies using resected brain tissues underscore the diagnostic and therapeutic potential of identifying somatic mutations, especially in drug-resistant focal epilepsy cases.^53^ Integrating germline and somatic testing into routine diagnostics could bridge this gap, facilitating a deeper understanding of lesional epilepsy and refining treatment strategies.^54^ The inclusion of such data alongside phenotypic and imaging insights offers a promising avenue for advancing precision medicine in epilepsy care.^55^

Our study has several limitations that nonetheless underscore critical avenues for future research and refinement. First, although ClinVar’s recently introduced Mondo Disease Ontology (Mondo) annotation system has enabled our large-scale, ontology-driven filtering of epilepsy-related variants, only about half of all ClinVar entries currently contain Mondo terms. This incomplete annotation may overlook a proportion of clinically and genetically relevant submissions. Yet, our method represents the largest and most comprehensive effort to date to harmonize Mondo-derived epilepsy terms with the ILAE Classification, demonstrating the scalability of ontology-based approaches for large-scale variant repositories. Second, while the ILAE Classification provides a clinically robust framework that aligns with widely accepted care standards, its age-dependent focus may inadvertently exclude important genes associated with epilepsies that do not map neatly into these categories—particularly malformations of cortical development (e.g., *FLNA*, *PAFAH1B1*, or *WDR45*) and neurodevelopmental disorders (NDDs) or autism spectrum disorders (ASD) with overlapping epileptogenic features. This constraint reflects the tension between following a standardized clinical categorization and acknowledging rapidly expanding genetic and phenotypic landscapes in epilepsy and related neurodevelopmental conditions. A dynamic process of expert curation and regular ontology updates will be paramount to integrate newly validated epilepsy genes and evolving phenotypes, especially those recognized in emerging diagnostic frameworks. Third, our conservative filtering criteria, including the requirement for at least one pathogenic or likely pathogenic variant in each gene, provided high-confidence associations but may have removed genes or variants that are still under investigation. This approach safeguards data quality yet may underestimate the number of potential epilepsy-related genes in early stages of validation. Incorporating curated but lower-evidence gene sets (e.g., those categorized as “moderate” or “limited” validity by ClinGen or identified in smaller case series) could offer a more comprehensive view of the evolving epilepsy genetic landscape. Fourth, our exclusive reliance on germline variants restricts the identification of somatic mutations implicated in lesional epilepsies, such as focal cortical dysplasia (FCD) and hemimegalencephaly, which frequently involve brain-tissue-specific variants in *DEPDC5*, *TSC1*, and *TSC2*.^50,52,53^ Capturing these somatic variants in large databases like ClinVar remains challenging, but doing so will be essential to improve our understanding of lesional and non-lesional epilepsy subtypes, advance tissue-specific precision medicine, and provide a more accurate depiction of the true genetic scope of epilepsy. Fifth, despite ClinVar’s global remit, submissions are influenced by the availability of genetic testing and data-sharing policies, which often favor high-resource settings. Regions with limited testing accessibility are underrepresented, potentially skewing the resulting snapshot of epilepsy genetics. Ongoing efforts to promote more equitable data sharing and to expand Mondo annotations internationally could mitigate these biases and enhance the inclusivity of large-scale data repositories. Finally, although the ILAE Classification is periodically updated to incorporate evolving clinical and genetic insights, a lag remains between these revisions and practical implementation in large databases. As new epilepsy genes emerge and existing evidence strengthens or weakens, dynamic feedback from the community—along with integration of complementary ontologies such as HPO for more granular phenotypic details—will be necessary to refine genotype-phenotype associations. Together, these efforts will strengthen the foundation for precision medicine in epilepsy, extending beyond infancy- and childhood-onset disorders to capture the full complexity of age-dependent and variable-age epilepsies, as well as overlapping neurodevelopmental phenotypes. Despite these limitations, our study lays the groundwork for systematically harnessing Mondo, the ILAE Classification, and expert curation frameworks to fuel ongoing discoveries and improve clinical translation in epilepsy genetics.

In summary, we demonstrate how leveraging the Mondo ontology for epilepsy-directed filtering of ClinVar data can systematically uncover and characterize ILAE-defined, age-dependent epilepsy syndromes. This scalable approach is readily adaptable to other epilepsy subtypes and broader human disease domains, bridging large genomic databases with clinically actionable insights. The resulting dataset serves as a valuable resource for guiding clinicians in the management of individuals across diverse age groups, facilitating specialized clinic development, and improving patient triage for targeted genetic care. It also supports clinical laboratories in designing refined gene panels and aids researchers in prioritizing candidate genes. Finally, these findings can inform expert stakeholders in formulating future guidelines and curation initiatives, ultimately propelling precision medicine in epilepsy and beyond.

## Supporting information

Supplemental Methods and Results

## Data Availability

Data produced in the present study are available in the manuscript text and supplemental files. Any additional data is available upon reasonable request to the authors.

