## Supplemental Methods and Results for "Harnessing Mondo and the ILAE Classification for Curation and Analysis of 71,942 epilepsy patient variants"

**Supplement**

### Supplemental Methods

#### Gene list curation

The ClinGen gene-disease validity data set was downloaded from the ClinGen website on January 7^th^, 2024. The data set provided gene symbol and HUGO Gene Nomenclature Committee Identifier (HGNC ID) for gene identification and MONDO disease label and ID for phenotype identification. We filtered the initial data set of 2,398 curations for the epilepsy genes identified through epilepsy-related MONDO terms and manually reviewed the resulting curations to resolve the possible discrepancies between the phenotypes curated by ClinGen and phenotypes logged in ClinVar for particular genes. We performed an expert annotation of gene variant-phenotype pairs in ClinVar as follows:

I. “Has gene-disease validity”: if the MONDO term for the submitted variant corresponded to the ClinGen curation for the particular gene and the evidence for validity was “Limited” and higher;
II. “No gene-disease validity”: if the evidence for validity was “Disputed” or “Refuted” or if a submitted MONDO term denoted a specific syndrome that corresponds to another gene (e.g., “alpha-methylacyl-CoA racemase deficiency” (MONDO:0013681) specific for disruptions in *AMACR* gene submitted for *POLG*);
III. “Not curated”: gene for the submitted variant is not in the ClinGen database, or non-curated phenotype for a gene where other phenotypes have been curated.

The complexity of phenotype associated with a curated gene might have prompted the ClinGen working group to assign a broad phenotype category associated with a gene, which could have several correlates in the phenotypes submitted to ClinVar. For example, the epilepsy phenotype associated with *SCN2A* is curated under the MONDO term “Complex neurodevelopmental disorder” (MONDO:0100038), while no *SCN2A* ClinVar entries feature this term. For cases of such disagreement, we reviewed the curation description and listed literature to derive MONDO terms that correspond to the curated phenotype. In case with the described example of *SCN2A* phenotype, “developmental and epileptic encephalopathy” (MONDO:0100062) and more specific term “developmental and epileptic encephalopathy, 11” (MONDO:0013388) were annotated as “Has gene-disease validity”, “seizures, benign familial infantile, 3” (MONDO:0011904) were annotated as “Not curated” as the working group specified this phenotype will be curated separately, and “neuronal ceroid lipofuscinosis 8” (which is associated with variants in *CNL8*) was annotated as “No gene-disease validity”.

#### International League Against Epilepsy (ILAE)-defined proxy age-dependent syndromes used to define specific syndromes

ILAE’s Classification of Epilepsy Syndromes summarized age-dependent epilepsies on the syndromic level, whereas ClinVar entries are oftentimes annotated with a single specific term that describes the disease. For example, phenotypes associated with *TSC1* entries are annotated as tuberous sclerosis 1 (MONDO:0008612). While tuberous sclerosis is one of the most common causes of infantile spasms and West syndrome, which are classified by ILAE as a syndrome with neonatal/infantile onset, tuberous sclerosis itself is not denoted with any specific onset. We identified such discrepancies across our gene set and consulted ILAE Classifications of Epilepsy Syndromes^1–4^ and a list of essential epilepsy genes available at epilepsydiagnosis.org. We re-annotated discrepant genes mentioned by the ILAE classifications as associated with a particular age-specific syndrome. The table lists reclassified genes and phenotypes.

| Gene | ClinVar MONDO term | ILAE phenotype proxy MONDO term | Age-related classification |
| --- | --- | --- | --- |
| *TSC1* | tuberous sclerosis 1 MONDO:0008612 | West syndrome (MONDO:0018097) | Neonatal/infantile |
| *TSC2* | tuberous sclerosis 2 (MONDO:0013199) | West syndrome (MONDO:0018097) | Neonatal/infantile |
| *FOXG1* | FOXG1 disorder (MONDO:0100040); Rett syndrome, congenital variant (MONDO:0013270) | West syndrome (MONDO:0018097) | Neonatal/infantile |

### Supplemental Figures

**Figure S1.** **A.** Mondo is a comprehensive, hierarchical disease ontology that is semi-automatically constructed from multiple disease-oriented ontologies. Its hierarchical structure allows for filtering and exploration at varying levels of disease term specificity, with "parent" terms representing broader categories and "child" terms offering more specific designations. In ClinVar, most submitted variants are annotated with Mondo terms to denote associated phenotypes, which facilitates automatic processing and disease-related subsetting of the data, thereby streamlining the analysis of genetic variants in a clinically relevant context. **B.** Challenges in using ClinVar for disease-oriented research and their solutions using Mondo ontology.

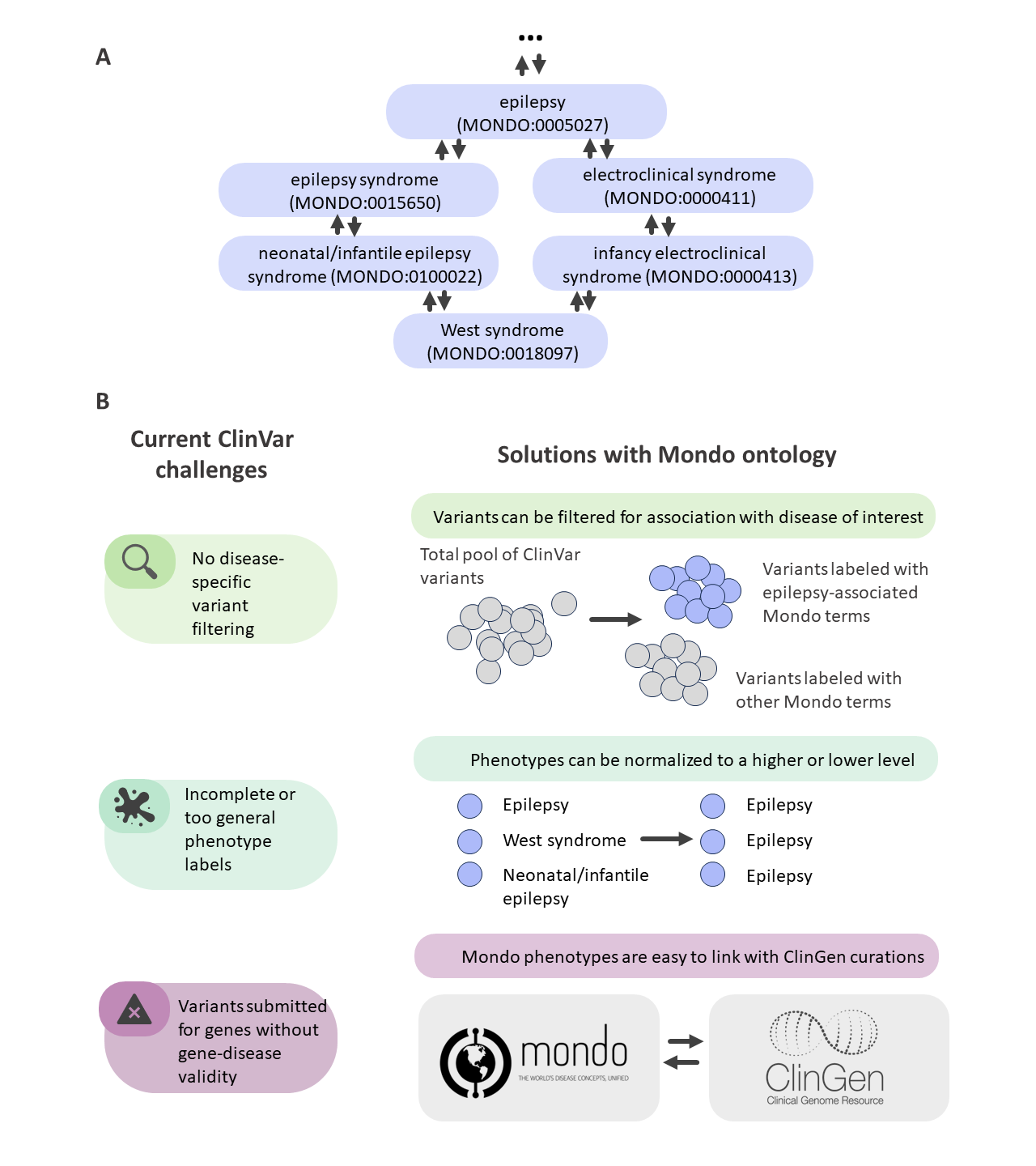

**Figure S2.** Proportion of pathogenic variants, VUS, and benign variants (A) and ratio of pathogenic variants to VUS (B) across the gene list levels (92 genes of level 1, 36 genes of Level 2, and 60 genes of Level 3). Level 3 genes had a higher proportion of pathogenic variants and lesser proportion of benign variants than genes of Level 1 and Level 2, whereas the proportion of VUS was similar across the three levels.

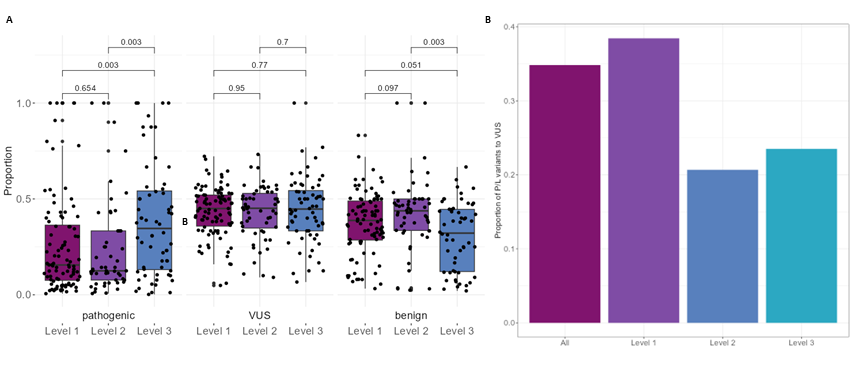

### Supplemental Results

Two genes that were not included in the Level 3 (i.e., not featured in any expert-curated effort) were *ARSD* (with two pathogenic variants associated with progressive myoclonic epilepsy and myoclonic astatic epilepsy, respectively) and *SLC28A2* (with two pathogenic variants associated with developmental and epileptic encephalopathy and one variant associated with progressive myoclonic epilepsy). Literature review did not identify any molecular, preclinical, or clinical evidence of association of either *ARSD* or *SLC28A2* with seizures or epilepsy. Thus, both genes and associated variants were excluded from the further analysis.

To explore the possibility of bias towards the genes that have been described earlier, we repeated the analysis outlined in the main results only using the variants submitted to ClinVar in the last two years from the ClinVar data acquisition (February 29, 2024). Filtering the age-dependent epilepsy syndrome dataset resulted in a set of 159 genes with 6,761 unique and 6,867 patient submissions of VUS and pathogenic variants.

Four genes that were present in the list of 50 genes with most VUS and pathogenic/likely pathogenic (P/LP) variants did not have submissions of new pathogenic variants in the past two years (*PNPO, CLN3, CLN5, CACNA1E*).

Figure S3 shows 50 genes with the highest number of pathogenic variants sorted by the total patient variant submissions similar to Figure 2 in the main results. The composition of the top 50 genes list is similar in the full data set and in the dataset with submissions for the past two years, with differences in the ranks highlighted in Figure S4.

Figure S5 features genes and phenotypes with the most submitted variants for the past two years per age category. Notably, the absolute numbers of submissions and gene ranks per category submitted in the past two years highly correlated with the number of all-time submissions for neonatal-onset (r=0.81 (95%CI 0.75-0.86), p < 2.2e-16) and childhood-onset (r=0.87 (95%CI 0.73-0.94), p= 5.012e-09) epilepsy syndromes and weakly for epilepsy syndromes with variable onset (r=0.37 (95%CI 0.12-0.57), p=0.004).

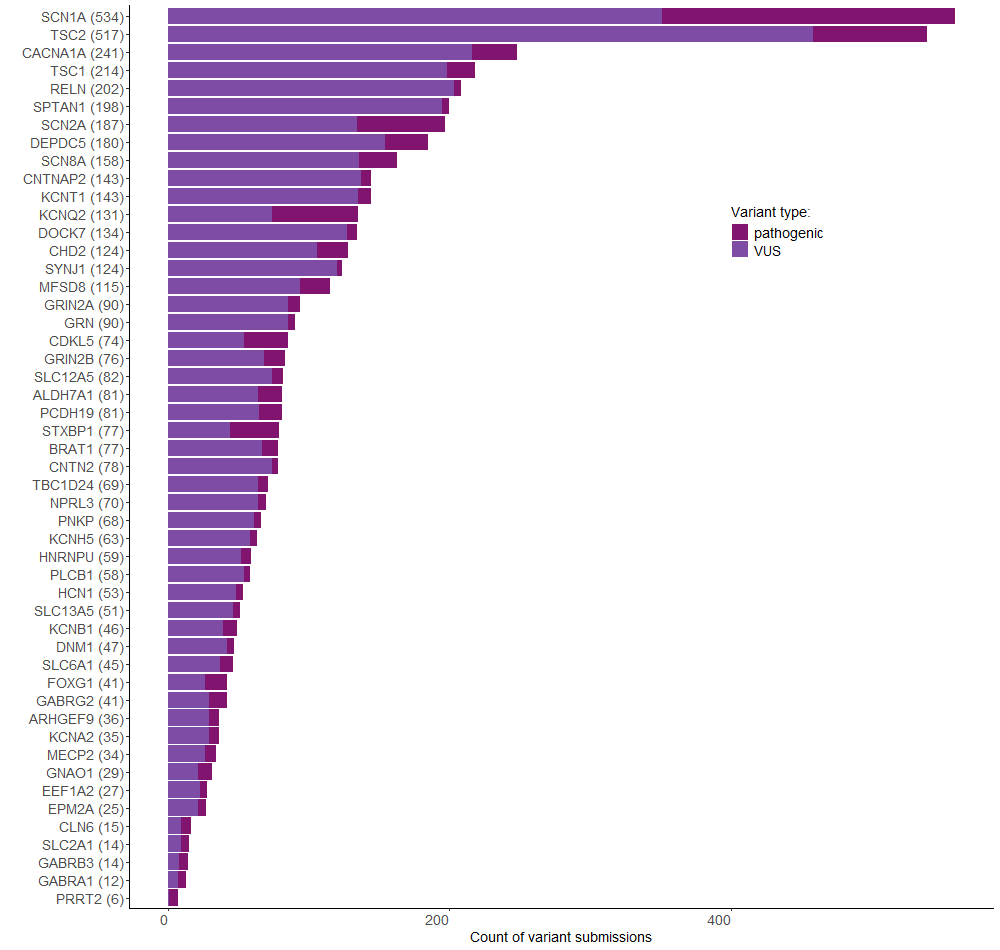

**Figure S3.** Fifty MONDO-defined epilepsy genes with the highest number of VUS and pathogenic variant patient submissions submitted in the last two years. The numbers in brackets next to the gene names indicate the total number of submissions, and the color designates the clinical significance of the variants.

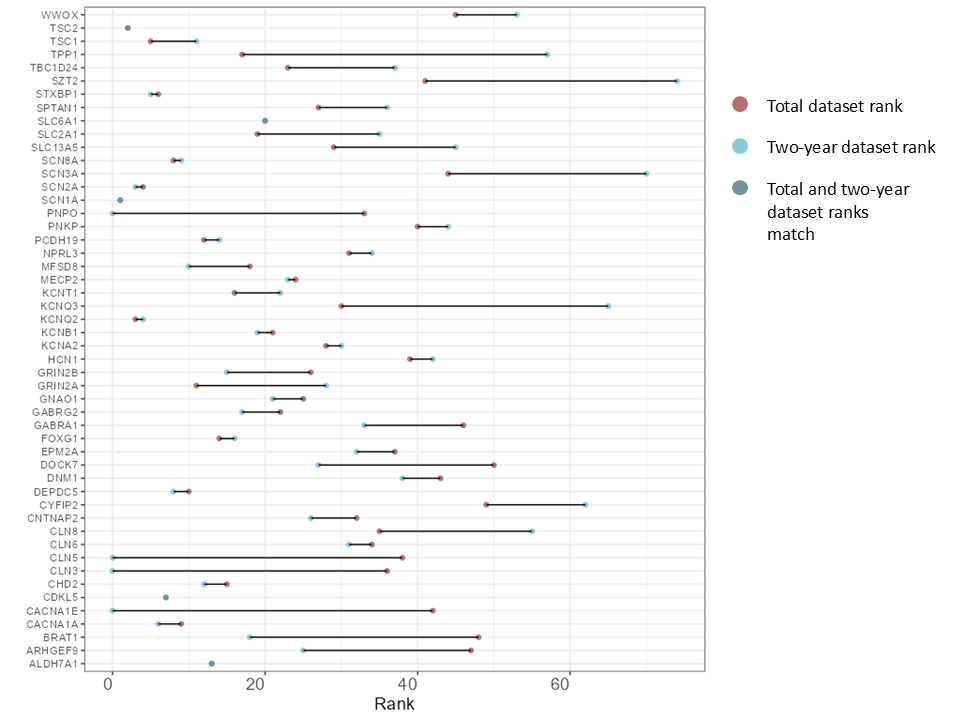

**Figure S4.** Change in ranks of 50 genes with the highest number of pathogenic variant patient submissions in the full dataset and dataset of patient submissions for the past two years. Four genes that were present in the list of 50 genes with most VUS and pathogenic/likely pathogenic (P/LP) variants did not have submissions of new pathogenic variants in the past two years (*PNPO, CLN3, CLN5, CACNA1E*).

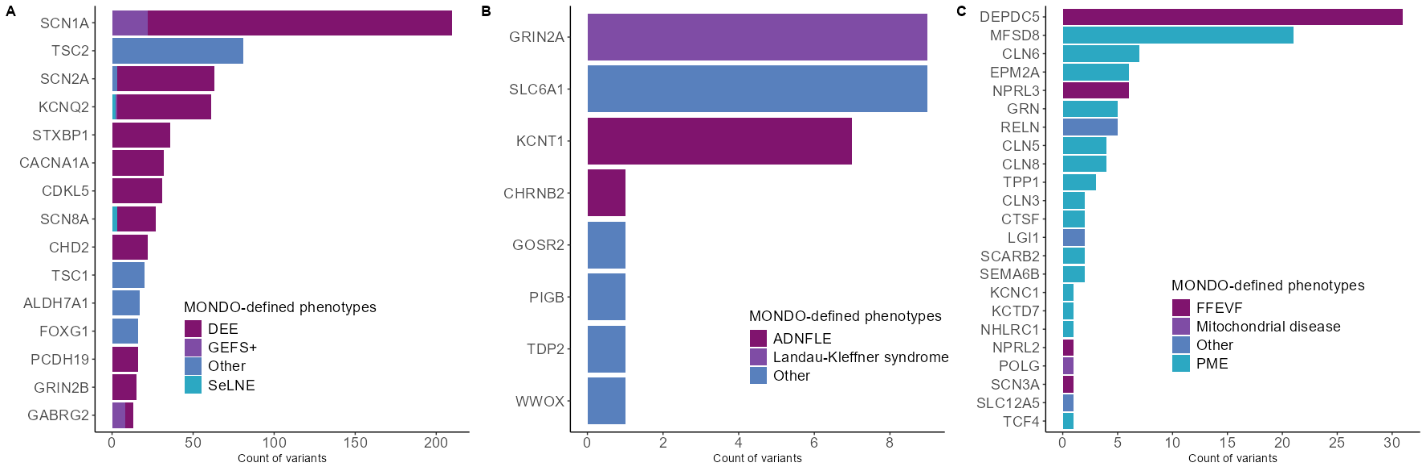

**Figure S5.** Bar plots showing pathogenic variant submission counts for the last two years for genes with the highest number of pathogenic variant submissions associated with MONDO-defined epilepsy syndromes with neonatal/infantile (B), childhood (C), adolescent/adult (D), and variable (E) onset. Only genes with more than 5 pathogenic variants submissions are displayed. The color codes for the MONDO phenotypes. Number- or gene-denoted phenotypes were aggregated into higher (more unspecific) MONDO hierarchy (for example, specific MONDO terms like developmental and epileptic encephalopathy, 42 (MONDO:0014917) and CDKL5 disorder (MONDO:0100039) were aggregated into developmental and epileptic encephalopathy (MONDO:0100062)).

Summary of variant submissions across all age-dependent groups in the dataset including variants submitted in the last two years.

| label | levels | Neonatal/infantile onset | Childhood onset | Variable onset | p |
| --- | --- | --- | --- | --- | --- |
| clinical_significance | benign | 4179 (44.1) | 411 (49.3) | 757 (40.3) | <0.001 |
|  | VUS | 4395 (46.3) | 392 (47.1) | 1011 (53.8) |  |
|  | pathogenic | 912 (9.6) | 30 (3.6) | 110 (5.9) |  |
| consequence_plot | Missense | 4838 (51.0) | 429 (51.5) | 1066 (56.8) | <0.001 |
|  | PTV | 760 (8.0) | 53 (6.4) | 105 (5.6) |  |
|  | Synonymous | 2015 (21.2) | 219 (26.3) | 339 (18.1) |  |
|  | intronic | 1519 (16.0) | 131 (15.7) | 211 (11.2) |  |
|  | Other | 354 (3.7) | 1 (0.1) | 157 (8.4) |  |
| gene_dis_val | yes | 7240 (76.3) | 370 (44.4) | 1161 (61.8) | <0.001 |
|  | Not curated | 2246 (23.7) | 463 (55.6) | 717 (38.2) |  |
